# SARS-CoV-2 re-infection risk in Austria

**DOI:** 10.1101/2021.02.08.21251362

**Authors:** Stefan Pilz, Ali Chakeri, John PA Ioannidis, Lukas Richter, Verena Theiler-Schwetz, Christian Trummer, Robert Krause, Franz Allerberger

**Author notes:** Contributed equally as first authors to this manuscript (Stefan Pilz, Ali Chakeri). **Corresponding Author**: Stefan Pilz, MD, PhD; Medical University of Graz; Department of Internal Medicine, Division of Endocrinology and Diabetology; Auenbruggerplatz 15, 8036 Graz, Austria. **Funding:** There was no specific funding for this work. **Conflicts of interest:** The authors report no conflicts of interest. **Contributions:** All authors have substantially contributed to the design, performance, analysis and reporting of the work. AC, LR and FA contributed to data collection. SP and JPAI analyzed the data and wrote the manuscript.

## Abstract

**Background:** A key question concerning coronavirus disease 2019 (COVID-19) is how effective and long lasting immunity against this disease is in individuals who were previously infected with severe acute respiratory syndrome coronavirus 2 (SARS-CoV-2). We aimed to evaluate the risk of SARS-CoV-2 re-infections in the general population in Austria.

**Methods:** This is a retrospective observational study using national SARS-CoV-2 infection data from the Austrian epidemiological reporting system. As the primary outcome, we aim to compare the odds of SARS-CoV-2 re-infections of COVID-19 survivors of the first wave (February to April 30, 2020) versus the odds of first infections in the remainder general population by tracking polymerase chain reaction (PCR)-confirmed infections of both groups during the second wave from September 1 to November 30, 2020. Re-infection counts are tentative, since it cannot be excluded that the positive PCR in the first and/or second wave might have been a false positive.

**Results:** We recorded 40 tentative re-infections in 14,840 COVID-19 survivors of the first wave (0.27%) and 253,581 infections in 8,885,640 individuals of the remaining general population (2.85%) translating into an odds ratio (95% confidence interval) of 0.09 (0.07 to 0.13).

**Conclusions:** We observed a relatively low re-infection rate of SARS-CoV-2 in Austria. Protection against SARS-CoV-2 after natural infection is comparable to the highest available estimates on vaccine efficacies. Further well-designed research on this issue is urgently needed for improving evidence-based decisions on public health measures and vaccination strategies.

## Introduction

The coronavirus disease 2019 (COVID-19) pandemic is a major public health crisis.^1,2^ A key question concerning measures against COVID-19 is the strength and durability of immunity against this disease in individuals previously infected with severe acute respiratory syndrome coronavirus 2 (SARS-CoV-2).^3-10^ Vaccination strategies, considerations regarding herd immunity, and overall simulations for the pandemic depend on the efficacy and the time course of immunity against COVID-19.^5^

Data on immune responses to COVID-19 are limited by knowledge gaps regarding their dynamics over time and their clinical significance with reference to protection against re-infections.^3-10^ There is evidence for re-infections from numerous case reports, but it is occasionally challenging to differentiate true re-infections from prolonged viral shedding that may last for up to about 4 months.^5,11,12^ Notably, a study of 12,541 health care workers in the UK recently found major protection against re-infection for those who had anti-SARS-CoV-2 antibodies determined by anti-spike and anti-nucleocapsid assays versus those who did not.^13^ After a follow-up of up to 31 weeks, they calculated a rate ratio of 0.11 (95% confidence interval (CI): 0.03 to 0.44; p=0.002) for re-infections in seropositive healthcare workers versus first infections in health care workers with negative antibody status.^13^ Similarly, another recent study among health care workers from the UK reported no re-infection case in 1038 individuals with evidence of previous SARS-CoV-2 infection based on PCR tests and/or antibody status.^10^ While these studies suggest a high protection against SARS-CoV-2 re-infections in health care workers, the risk of re-infections in the general population remains uncertain.

Austria was hit very early in this pandemic with a first wave occurring from February 22 to April 30, 2020 (all further dates refer to the year 2020). Data on the re-infection rate during the second wave from September 1 to November 30, can therefore provide, as a rough estimate, evidence on the immunity against SARS-CoV-2 over more than half a year.^14,15^ Therefore, we investigated data from the Austrian epidemiological reporting system (ERS) provided by the Austrian Agency for Health and Food Safety (AGES).^15^ As the primary outcome, we compared the odds for SARS-CoV-2 re-infections in COVID-19 survivors versus first infections in the remainder general population during the second infection wave. In addition, we also evaluate data on hospitalization status during both infection waves and on COVID-19 deaths during the second wave, in order to obtain measures of disease severity.

## Methods

Data for this study were derived from the Austrian ERS that is tracking SARS-CoV-2 infection data in Austria, including amongst others data on hospitalization status and COVID-19 deaths.^15^ Ethical approval for this study was obtained from the ethics committee at the Medical University of Graz, Graz, Austria.

Patients who had a positive polymerase chain reaction (PCR) test during both, the first and second infection wave are referred to here as patients with “tentative re-infections”. We use the term “tentative” re-infection because a certain number of these cases might reflect false-positive results in the testing during the first and/or second wave. This is based on the consideration that the specificity (with 95% confidence region) of PCR tests (nucleic acid amplification tests) for SARS-CoV-2 is less than 100%, with 98.1% (95.9 to 99.2%) according to a recent meta-analysis.^16^

The group size of “COVID-19 survivors” was calculated as all individuals who had a positive PCR test result for SARS-CoV-2 minus all reported COVID-19 deaths from February 22 to April 30. The control group (“general population group”) are the remainder Austrian residents that we calculated as the reported Austrian population on January 1 with 8,901,064 individuals (the closest approximation for the population size) minus all patients tested SARS-CoV-2 positive during the first wave.^17^ In Austria, population changes from year to year are usually significantly less than 1%.^17^ The observation period for tracking SARS-CoV-2 infections was from September 1 to November 30 (the pre-specified date for our analyses), corresponding to what we term the second wave. Automated matching of records in the first and second wave to detect tentative re-infections was done by using IDs consisting of the first two initials of the first name, the first three initials of the surname and the date of birth (e.g. ST.PIL.15.12.1979). All entries with the identical ID were then carefully and manually checked including data such as full names and laboratory dates to evaluate whether the criteria for a re-infection were met.

We did not primarily track tentative re-infections of COVID-19 survivors from May to August as it may be unclear whether positive SARS-CoV-2 tests represented re-infection or persistent infection when considering long-term viral shedding for up to about 4 months.^5-7^ This 4 month interval was also the main consideration to separate the time frame for the two waves. Of note, there were only relatively few documented SARS-CoV-2 cases (<0.15% of the Austrian population) from May to August.^15^

Regardless of the main reason for hospitalization, any hospitalized patient who was tested SARS-CoV-2 positive was classified as hospitalized in the ERS. All persons who were tested SARS-CoV-2 positive and died for whatever reason within 28 days after the last positive test were classified as COVID-19 deaths.

As our primary outcome analysis, we calculated the odds ratio (OR) (with 95% confidence interval [CI]) of SARS-CoV-2 re-infections in the COVID-19 survivor group versus first infections in the general population group (see Figure 1 for our analysis plan). Statistical analyses were performed by using SPSS Version 25.0 (IBM SPSS Inc., Chicago, IL, USA).

**Figure 1.**
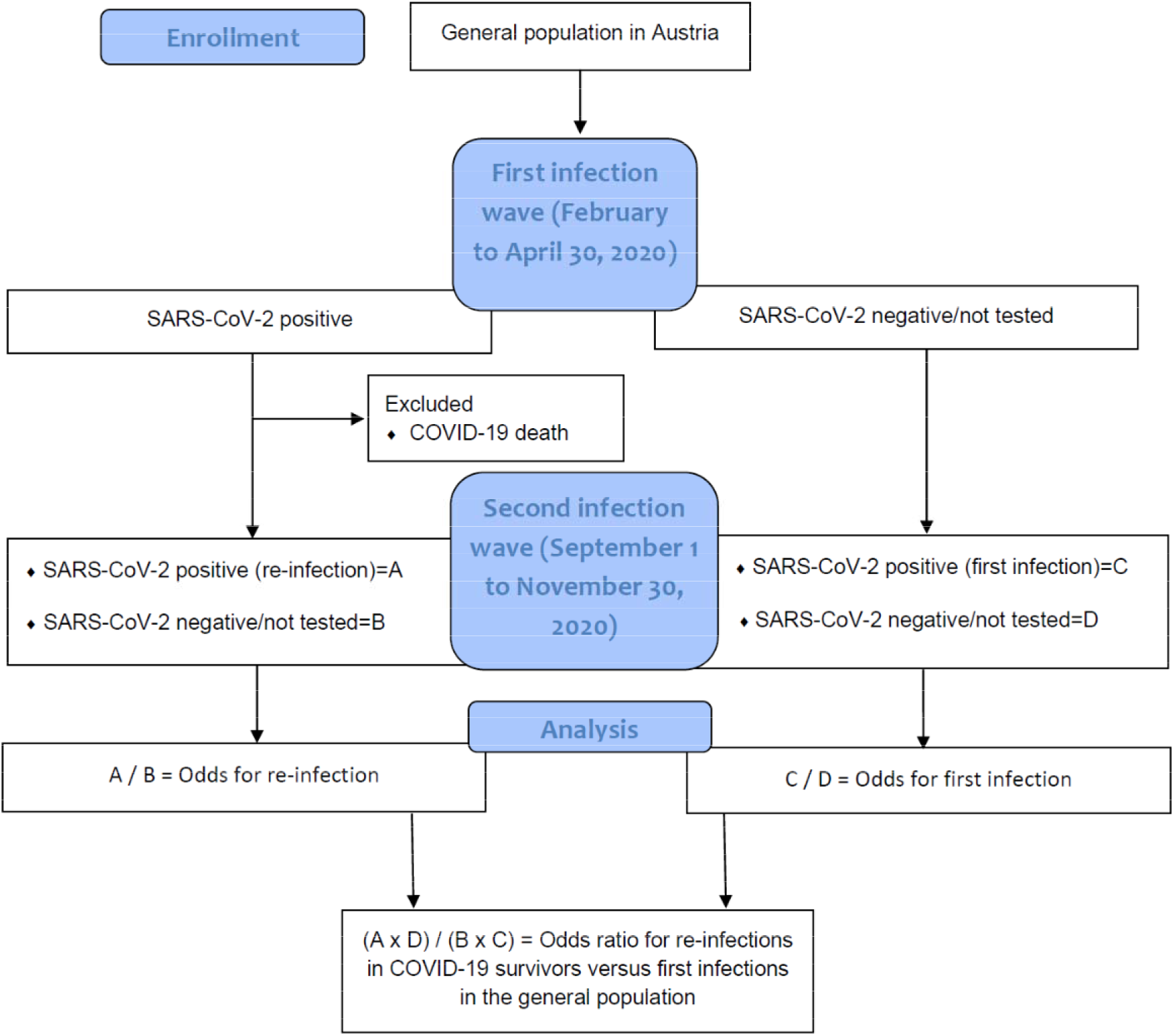
Flow chart and analysis plan.

## Results

From 15,424 patients with SARS-CoV-2 positive tests in the first wave, 584 were recorded as COVID-19 deaths, so that our COVID-19 survivor group consists of 14,840 patients. Excluding the COVID-19 survivor group, the number of individuals of the general population group resulted in 8,885,640 individuals.

During the observation period from September 1 to November 30, we recorded 40 tentative re-infections in the COVID-19 survivor group (0.27%), and 253,581 new infections in the general population group (2.85%). The OR (with 95% CI) for infections in the COVID-19 survivor group versus the general population group was 0.09 (95% CI, 0.07 to 0.13).

Characteristics of the 40 re-infection cases are tabulated in Table 1. Of the patients with tentative reinfections, 62.5% were women and the median age (with 25^th^ to 75^th^ percentile; minimum – maximum) at the first infection was 39.8 (25.9 to 54.5; 15.4 – 93.8) years. The mean (± standard deviation) time from the first to the tentative re-infection was 212 ± 25 days. Of the 40 tentative re-infections, 4, 12, and 24 were documented in September, October, and November, respectively (among 18,106, 61,384, and 174,131 total infections, respectively).

**Table 1.** Characteristics of 40 patients with re-infection

| Gender | Age at first infection(years) | Time between infections (days) | Hospitalization |  |
| --- | --- | --- | --- | --- |
|  |  |  | First wave | Second wave |
| Female | 80-85 | 148 | Yes | No |
| Female | 50-55 | 223 | No | No |
| Female | 50-55 | 183 | No | No |
| Male | 30-35 | 215 | Yes | No |
| Female | 30-35 | 200 | Unknown | Unknown |
| Female | 25-30 | 206 | No | No |
| Male | 85-90 | 196 | No | No |
| Female | 35-40 | 175 | No | No |
| Male | 50-55 | 222 | No | Unknown |
| Male | 20-25 | 251 | No | Unknown |
| Female | 80-85 | 148 | Yes | Yes |
| Male | 75-80 | 238 | Yes | Unknown |
| Female | 20-25 | 236 | No | Unknown |
| Female | 55-60 | 214 | No | No |
| Female | 35-40 | 203 | No | No |
| Female | 20-25 | 222 | No | No |
| Male | 15-20 | 235 | No | No |
| Female | 75-80 | 219 | Yes | Yes |
| Male | 50-55 | 206 | No | No |
| Female | 70-75 | 172 | No | No |
| Male | 20-25 | 207 | No | No |
| Female | 50-55 | 221 | No | No |
| Male | 15-20 | 210 | No | No |
| Female | 40-45 | 246 | No | No |
| Male | 60-65 | 246 | No | Unknown |
| Male | 25-30 | 221 | Yes | Yes |
| Male | 45-50 | 232 | No | No |
| Female | 30-35 | 222 | No | Yes |
| Female | 30-35 | 231 | No | No |
| Female | 30-35 | 213 | No | No |
| Female | 50-55 | 173 | Yes | Yes |
| Male | 25-30 | 203 | No | No |
| Female | 20-25 | 172 | No | No |
| Female | 40-45 | 214 | No | Unknown |
| Male | 25-30 | 221 | No | No |
| Female | 90-95 | 237 | Yes | Unknown |
| Female | 25-30 | 227 | No | No |
| Female | 40-45 | 226 | No | No |
| Female | 45-50 | 216 | No | No |
| Male | 25-30 | 243 | No | No |

Hospitalization status in numbers of patients coded as yes, no and unknown was 8, 31 and 1 for the first infection and 5, 27 and 8 for the tentative re-infection, respectively. Four patients were hospitalized during both infection waves. Unknown hospitalization data during the second wave are probably mainly due to a delay in hospitalization data entry into the ERS.

With follow-up on mortality available until December 23, only one woman in the age range 70 to 75 years died two days after her tentative re-infection diagnosis. She was not hospitalized and according to her medical records her cause of death (“acute vascular occlusion of an extremity with rhabdomyolysis”) was not causally attributed to COVID-19.

## Discussion

We documented a relatively low re-infection risk for SARS-CoV-2 in the general population of Austria by using data from the ERS. Patients with re-infections covered both genders, a wide age range and included also patients who were hospitalized during both infections.

Our study is, to the best of our knowledge, the first systematic investigation of tentative re-infection risk with SARS-CoV-2 in a large national population. Several case reports on SARS-CoV-2 re-infections in the general population indicate that there is at least some risk of re-infection, but they did not provide quantification of re-infection risk that requires a standardized comparison to the “background” infection risk in the general population.^3-5^ While data on immune responses to previous SARS-CoV-2 infections exist, they can only be regarded a proxy for a previous infection and the associated clinical protection against re-infections, thus requiring studies like ours to address the question to what extent patients who experienced PCR confirmed SARS-CoV-2 infections are protected against re-infections.^3-5^ Importantly, the study by Lumley et al in 12,541 healthcare workers documented protection against re-infection for those who had anti-SARS-CoV-2 antibodies with a rate ratio (0.11) very similar to what we observed.^13^ While the investigation by Lumley et al was restricted to a specific population of predominantly healthy adult health care workers 65 years of age or younger, and was based on only two re-infections in seronegative individuals, our study extends this knowledge by data from a much larger population based survey using solely PCR-confirmed SARS-CoV-2 infection cases.^13^ Importantly, a recent study using SARS-CoV-2 PCR and antibody test data from 66,001 patients from a laboratory in south-west London documented 8 patients with evidence of re-infections, and calculated a relative risk of re-infections versus first infections of 0.0578 (95% CI: 0.0288 to 0.1160)^18^ which is also compatible with our estimate.

Our data do not include detailed clinical characteristics of the patients with tentative re-infections but it is noteworthy that these patients covered both genders with a wide age range and included also several hospitalized patients. These data are of interest since previous studies indicate a high correlation between neutralizing antibodies against SARS-CoV-2 and COVID-19 severity. This in turn suggests that those patients with more severe infections may develop a stronger protective humoral immune response against SARS-CoV-2 compared to those with less severe infections. This hypothesis is, however, not strongly supported by our findings as several patients with tentative re-infections were already hospitalized during their first infection.^8^ Regarding duration of acquired immunity against SARS-CoV-2 re-infections, we provide data with a median follow-up time of about 7 months. Importantly, there was no clear sign of decreasing protection against re-infections in descriptive analyses of monthly stratified re-infection cases.

In view of ongoing discussions on vaccination approaches regarding SARS-CoV-2, our data suggest that the protection against SARS-CoV-2 after natural infection is roughly similar to the highest estimates of SARS-CoV-2 vaccine efficacies among vaccines that have been authorized to-date, although a direct comparison cannot be made due to differences in study designs and study populations.^19,20^Nevertheless, we believe that based on our findings, waving urgent recommendations to undergo SARS-Cov-2-vaccination for persons with PCR-documented previous COVID-19 infection seems prudent as long as any shortage of vaccines is present.

Our findings on a significant protection against SARS-CoV-2 re-infections, provide also evidence for the rapid evolution of the pandemic towards “herd immunity”, in particular because of a huge underreporting of SARS-CoV-2 cases.^21,22^ Therefore, the relatively high prevalence of individuals who were already infected with SARS-CoV-2 along with the currently rapidly increasing number of vaccinated individuals may work in concert towards an ensuring “herd immunity” that will hopefully bend this pandemic within the near future.^2,23,24^ This may already be the case in some countries such as India, where seroprevalence rose rapidly from 0.7% in May to 7% in August and 60% in November in national surveys.^25-27^ Accordingly, the epidemic wave in India (both for documented cases and for COVID-19 deaths) has largely abated by February 2021. It must, however, be noted that the concept of herd immunity has recently been challenged by resurgence of COVID-19 in Manaus, Brazil, a region in which seroprevalence data suggested that about 76% of the population had been infected with SARS-CoV-2 by October 2020.^28^ It is unknown whether there was an error with over-estimation of the first wave seroprevalence, or the resurgence can be explained by the advent of a new strain (P1) that has a high propensity for re-infection. Careful monitoring for new strains and for their ability to evade existing natural immune responses and vaccine-induced immunity is needed.

Our findings are limited due to lack of detailed clinical characteristics, the observational nature of our study design, and the strong dependence on the data quality of the ERS. The 40 tentative re-infections have quite similar demographics to the totality of COVID-19 documented cases in Austria, but data are limited for meaningful formal comparisons.^9^ Data on hospitalizations are very sparse and hospitalization data during the second wave are missing for some participants, probably, due to a delay in reporting such data. Infections in the first wave are likely to have been far more common than the documented ones, so some of the general population controls may actually represent people already infected in the first wave. Moreover, the relative risk of re-infection may be over-estimated, if re-infection cases are artifacts of PCR false positives in either wave; and underestimated if people who were infected in the first wave were less likely to be tested in the second wave compared with other people having the same symptoms. In this context, Lumley et al. reported that seropositive health care workers attended asymptomatic screening less often than seronegative health care workers with a rate ratio of 0.76 (95% CI: 0.73 to 0.80), a finding that is similar compared to another study from the UK.^10,13^ Another limitation of our work is that we did not have access to viral sequencing data to compare first and re-infections, and it is not known how well our findings generalize to the re-infection risk concerning different genetic variants of SARS-CoV-2. Finally, we have to stress that our main findings are only a rough estimate of SARS-CoV-2 re-infection risk, requiring urgent confirmation in other populations and study settings.

In conclusion, we observed a relatively low tentative re-infection rate of SARS-CoV-2 in Austria that suggests a similar protection against SARS-CoV-2 infection compared to vaccine efficacies.^5,19,20^ These data may be useful for decisions on public health measures and vaccination strategies to fight the COVID-19 pandemic.^2,19,20,23,24^ Further studies are urgently needed to improve our knowledge on SARS-CoV-2 re-infection risk and its predisposing factors and clinical significance.

## Data Availability

All relevant study data are already included in our manuscript.

## Acknowledgment

We thank all persons providing and entering data for this work.

